# Evidence-Based Quality Scores for Rating Drug Products and Their Utility in Health Systems

**DOI:** 10.1101/2020.05.22.20110775

**Authors:** Arash Dabestani, Carl W. Bazil, Ryan C. Costantino, Erin Fox, Joe Graedon, Harry Lever, Robert Makuch, C. Michael White

## Abstract

The quality of drug products in the United States, which are largely produced overseas, has been a matter of growing concern.^1^ Buyers and payers of pharmaceuticals, whether they are health-systems, insurers, PBMs, pharmacies, physicians, or patients, have little to no visibility into any quality metrics for the manufacturers of drug products or the products themselves. A system of “quality scores” is proposed to enable health-systems and other purchasers and payers of medication to differentiate among drug products according to evidence-based metrics. Metrics influencing the quality scores described herein include both broadly applicable regulatory information and more drug-specific, third-party chemical analysis information. The aggregation of these metrics through a proposed set of rules results in numerical values on a 0-100 scale that may be further simplified into a red/yellow/green designation. The simplicity of such scores enables seamless integration into existing healthcare systems and an integration scheme is proposed. Using real-world data from currently on-market valsartan drug products, this proposed system generated a variety of quality scores for six major manufacturers. These scores were further evaluated according to their current market price showing no significant correlation between quality score and price. The implementation of drug quality scores at healthcare institutions in the United States and their potential utilization by regulators, could create a much-needed, market-driven incentive for pharmaceutical manufacturers to produce quality medications that would reduce drug shortages and improve public health.

## Introduction

As most of the United States’ complex drug supply chain has moved overseas, especially to countries such as India and China, quality and safety concerns have become more pressing. Eighty percent of active pharmaceutical ingredients (“API”) for products sold in the U.S. now come from outside the country, the vast majority from China.^2^ As Dr. Janet Woodcock, Director of the Food and Drug Administration (“FDA”) Center for Drug Evaluation and Research (“CDER”), has noted, this “use of foreign-sourced materials creates vulnerabilities in the U.S. drug supply.”^3^ Recent drug quality issues have threatened the health and safety of American consumers, including the widespread contamination of critical blood pressure medications, ^4^ gastroesophageal reflux disease drugs,^5^ and diabetes medications^6^ with carcinogens.^7^ Not only do drug quality issues place patients’ lives at risk, they also account for over 60% of drug shortages^8^ and generate fear and mistrust that is an important cause of medication non-adherence.^9^

Certain manufacturers have exhibited substantive quality issues and even engaged in data manipulation. This issue is highlighted by the record $500 million fine imposed on the generics manufacturer, Ranbaxy, after it pleaded guilty to failing to report its drugs did not meet specifications. The firm also made false statements to the FDA. Ranbaxy knowingly manufactured drugs that tested out-of-specification, had unknown impurities, and would not maintain their expected shelf life.^10^

Although significant attention is given to overseas manufacturers, American companies are not immune from quality issues. Numerous cases exist of serious quality problems affecting American consumers caused by poor manufacturing practices at facilities in the United States.^11^

For these reasons, we applaud the FDA’s recent recognition of the need for more transparency with regard to drug manufacturing.^12^ Recalls and FDA investigations have made clear that not all manufacturers are alike in their capacity to reliably produce high-quality pharmaceutical products. However, purchasers of pharmaceutical products – including drug distributors, pharmacies, and health systems – often have no reliable way to distinguish between high- and low-quality manufacturers or their drug products.

The FDA’s Task Force on Drug Shortages has endorsed the creation of a voluntary “rating system…. to inform purchasers, group purchasing organizations (GPOs) for health care systems, and even consumers, about the quality management maturity of the facilities making the drugs.”^13^ This underscores the importance of the fundamental principle of having a quality score that can differentiate between manufacturers. However, since the FDA proposal is voluntary, it may not achieve broad implementation. Furthermore, it is important that the criteria used be evidence-based. Announcements have also been made by private industry for the creation of a commercially available drug quality scores platform intended for use by health systems.^14^

Any reliable rating system should draw upon objective, science-based, independently generated data that is not voluntarily provided by manufacturers but collected by independent parties. Although a quality score system may include voluntarily furnished data, it must be primarily based on independent data to be broadly applicable and thus optimally useful to healthcare systems. The American College of Cardiology stressed the need for “<u>independent</u> testing and verification of the chemical content of batches of pharmaceuticals” in a recent resolution^15^ that emphasizes the necessity to rely on more than just the manufacturer’s self-reported data.

These independent quality rating systems should be developed through a process that incorporates robust stakeholder feedback, including patients, providers, academic institutions, regulatory agencies and health systems. In order to spur such discussion and make meaningful progress towards establishing a viable system for use among an array of healthcare providers, the authors propose criteria for the creation of evidence-based quality scores, examples of use on existing drug products, and a mechanism for utilization exemplified by a proposed workflow for Health Systems.

## Methods

### Quality Score Overview

Evidence collected in this proposed system originates from both broad manufacturer-level data and from specific product information. The combination of this data is intended to influence scores for specific drug products of a particular drug from a specific manufacturer. Although the evidence can be aggregated to evaluate a given manufacturer as a whole, the greatest utility to healthcare purchasers and payers is likely achieved by focusing on specific products. This is due to the immense complexity and opacity of the pharmaceutical supply chain. The source of ingredients used in any one drug product is considered proprietary and is therefore not easily accessible.

The specificity down to a drug product is not intended to directly describe a given National Drug Code (“NDC”), which further defines a drug product’s dosage form and packaging. It is assumed that evidence gathered on a specific drug product will be applicable to all NDCs related to that drug product from the specific manufacturer, regardless of dosage level or packaging. As an illustrative example, if negative information is gathered for “manufacturer X’s” valsartan 160mg tablets packaged in 100 count bottles, this will influence quality scores on NDCs for all valsartan tablets in all package sizes for manufacturer X. When substantially more data is available, future iterations of quality scores may directly describe individual NDCs or individual dosage forms.

The proposed system would generate a quality score on a numerical scale from 0 to 100, with 100 being the most desirable and highest achievable score and 0 being the lowest and least desirable score. Since all drug products legally sold in the United States are FDA-approved and produced at registered facilities certified as conforming to Current Good Manufacturing Practices (“cGMP”), the default assumption is that, absent evidence to the contrary, all products receive a default score of 100.

Criteria proposed herein are all based on information that is negative in nature and thus produces evidence for reducing a starting score of 100. Future iterations of such quality scores may also include criteria based on positive information that generates evidence for raising a score. The default value of such scores may be subsequently lowered to add opportunity for particularly well-performing manufacturers or products to outperform the default. It is also contemplated that temporal considerations be given to modify the impact of negative information and to eventually remove or significantly reduce its influence. The intention for a reliable quality score system would be to continuously incorporate new regulatory and chemical analysis data to enable optimal, real-time, guidance of drug product quality.

### Quality Score Criteria

Proposed below are detailed criteria and their influence on a default score. These are based on independently gathered evidence from regulatory information and chemical analysis of on-market drug products obtained from a licensed pharmacy.

**Table 1.** Proposed quality score criteria are categorized by information derived from regulatory data and chemical analysis data. For most criteria, the severity of negative influence on the score is dependent on qualifiers on the information gathered.

| Category | Criteria | Qualifiers | Score Influence |
| --- | --- | --- | --- |
| Regulatory Information | Warning Letter ratio to total inspections | >1.5X 3-yr industry average<br>>2X 3-yr industry average | -10<br>-30 |
|  | Form 483 ratio to total inspections | >10% 3-yr industry average<br>>20% 3-yr industry average | -10<br>-30 |
|  | GMP related Consent Decree/ CIA in place |  | -50 |
|  | Public Product Quality complaints | e.g. % "bad odor" > 2X competitors<br>e.g. % "bad odor" > 4X competitors | -10<br>-30 |
|  | Serious Adverse event | e.g. % "death" > 2X competitors<br>e.g. % "death" > 4X competitors | -10<br>-30 |
| Chemical Analysis | Dosage failure | Single batch<br>>33% of batches<br>All batches | -10<br>-30<br>-61 |
|  | Dissolution failure of USP | Single batch<br>>33% of batches<br>All batches | -10<br>-30<br>-61 |
|  | Dissolution failure of Physiological Conditions | >33% of batches<br>All batches | -10<br>-30 |
|  | Carcinogen failure of FDA levels | Single batch<br>>33% of batches | -30<br>-61 |
|  | Carcinogen failure at evidence-based, stricter levels | Single batch<br>>33% of batches<br>All batches | -10<br>-30<br>-61 |
|  | Heavy metals failure of FDA levels | Single batch<br>>33% of batches | -30<br>-61 |
|  | Microbial detection failure by FDA method | Single batch<br>>33% of batches | -30<br>-61 |
|  | Microbial detection failure by PCR method | Single batch<br>>33% of batches<br>All batches | -10<br>-30<br>-61 |
|  | Ingredients ID failure, API | Single batch<br>>33% of batches | -30<br>-61 |
|  | Ingredients ID failure, excipient | Single batch<br>>33% of batches<br>All batches | -10<br>-30<br>-61 |

The specific criteria proposed above are primarily self-explanatory. Criteria requiring clarification are discussed below.

**Form 483 and Warning Letter Ratio of Inspections** – The 3-year average of total drug industry inspections, Form 483 letters and warning letters is aggregated and the ratios of Form 483 letters to total inspections and warning letters to total inspections is calculated. These same values are also calculated for an individual manufacturer and if the ratios for the manufacturer are higher than the global average by a set qualifier, a negative score influence is triggered. Future iterations may utilize total drug industry inspections within geographic regions as opposed to a global average. This could be an important refinement given the differences in inspection practices within the United States and overseas; such as domestic inspections are unannounced whereas foreign inspections often come with months of advanced warning.^16^

**Public Product Quality Complaints or Serious Adverse Events** – The ratio of this complaint or event to all others for this product is compared to other manufacturers of the same product. If the ratio for a concerning complaint or serious event is significantly higher than the average ratio of its competitors, a negative score influence is triggered.

**Dissolution Failure of Physiological Conditions** – This differs from dissolution failure of USP conditions for a variety of products where the registered USP monograph for dissolution testing does not conform to industry standard physiologically relevant conditions. For example, industry standard simulated gastric fluid is often used for 2 hours and has a pH of 1.2 and simulated intestinal fluid is often used for the remainder of dissolution testing thereafter and has a pH of 6.8. However, USP dissolution media for ibuprofen tablets prescribes using only one solution with a pH of 7.2 without any exposure to acid. Although testing ibuprofen tablets in USP solution may yield a passing test, performing dissolution testing in physiologically relevant media has been shown to yield certain specific products taking over 24 hours to dissolve whereas others dissolve quickly, as expected.^17^

**Carcinogen Failure at Evidence-based, Stricter Levels** – FDA regulations for acceptable daily exposures or intakes of various carcinogen compounds generally follow internationally accepted guidelines. However, there are cases where organizations such as the World Health Organization (“WHO”) and the International Agency for Research on Cancer (“IARC”) will provide guidance which differs from that listed by the FDA. This is currently the case with N,N-Dimethylformamide^18^ (“DMF”) which is classified by WHO and IARC as a Group 2A probable human carcinogen.^19^ For the purposes of this proposed quality score system, a negative score influence is triggered when DMF levels exceed 96 nanogram but are less than 1,000 nanograms and a more severe negative score influence is triggered when DMF levels exceed 1,000 nanograms.

**Table 2.** Quality score criteria definitions for “Carcinogen failure at evidence-based, stricter levels” specific for DMF

| Category | Criteria | Qualifiers | Score Influence |
| --- | --- | --- | --- |
| Chemical Analysis | Carcinogen failure: DMF >96 ng, < 1,000ng | >33% of batches | -10 |
|  |  | All batches | -30 |
|  | Carcinogen failure: DMF >1,000 ng | Single batch | -10 |
|  |  | >33% of batches | -30 |
|  |  | All batches | -61 |

Notably absent from the proposed quality score criteria is information regarding recalls. Although the existence of high volumes of recalls for a particular manufacturer of a drug product may intuitively induce a negative score influence, this may, in fact, be an indication of responsible quality surveillance. Furthermore, a lack of recalls may be indicative of overly lax quality assurance measures for a given manufacturer as opposed to a truly quality product. In the United States, drug product recalls are almost all voluntary and performed at the discretion of pharmaceutical manufacturers.^20^ This conundrum warrants a deeper investigation. A retroactive review of chemical data compared with recall data could potentially better inform the correct view of product recalls. While such insights are yet to be elucidated, it was deemed best to leave such information out of the currently proposed quality score system.

Also absent from the quality score criteria is the FDA-proposed concept of quality management maturity. Indicators of quality management maturity have been proposed but appear to primarily rely on manufacturers’ proprietary information.^21^ To the authors’ knowledge, there is no existing metric that uses publicly available inputs other than recalls which are discussed above. The lack of available information to assess the merits of quality management maturity for use in an independently derived and broadly applicable, evidence-based quality score system precludes it from inclusion in this proposal; however, future iterations may add such criteria when the information required for evaluation is made available or new indicators are elucidated.

It is envisioned that a drug quality score system or platform could include a mechanism for health system users to report potential drug quality issues, adverse events or send suspect medication samples for chemical analysis. This could create a much broader net to identify quality issues and if broadly utilized, such information could be valuable for the creation of new criteria to influence quality scores.

### Quality Score Mechanics

To enable further ease of use and straightforward implementation within established healthcare systems, the proposed numerical quality score output can be categorized in a red/yellow/green fashion according to the following table:

**Table 3.** Quality scores receive a color designation dependent on their numerical value.

| Color Designation | Quality Score Range |
| --- | --- |
| Green | 80-100 |
| Yellow | 40-79 |
| Red | 0-39 |

Recognizing that a drug product receiving a red designation could induce significant impact within a healthcare system; special consideration was given to criteria which can trigger a red. In this proposal, only the quality score criteria within the category of Chemical Analysis is allowed to trigger a red designation. Even if the sum of Regulatory Information criteria resulted in a score influence of -61 or below, the reported quality score would be a minimum of 40, yielding a yellow designation. The logic for this is rooted in the assumption that regulatory findings and public reporting can be influenced by many factors and do not have a well-established correlation to product quality, which is defined by its chemical composition. Supporting this is an excerpt from a 2015 White Paper from the FDA Office of Pharmaceutical Quality:^22^

> “FDA has only limited information about the current state of pharmaceutical quality. FDA has no formal means for quality surveillance, except through inspections
>
> …
>
> Furthermore, inspection findings have not been a reliable predictor of the state of quality.”

### Proposed Implementation for Health Systems

The intended use of the proposed quality scores system in an established healthcare system would be to inform and enable pharmacy procurement teams so that decision trees could be enacted. Decision trees could be implemented through healthcare IT systems that standalone or are integrated into the health systems’ existing vender or purchasing system. A proposal of a decision tree utilizing such quality scores in order to purchase primarily green, occasionally yellow after manager review and completely avoid red is proposed for a health system where a robust process exists for managing drug shortages. Such drug shortage processes may include identification of substitute products, determination of alternative drugs or treatments and other remedies for mitigating or minimizing the impact of a drug shortage. In extreme cases that are reviewed by management, a poorly scoring medication product where there is no alternative could be treated by the health system as a drug shortage instead of purchasing a product designated red. Depending on the healthcare system, it may require a different decision tree and may elect to utilize different criteria, or adopt the same criteria with different degrees of influence on the quality score values.

**Figure 1.**
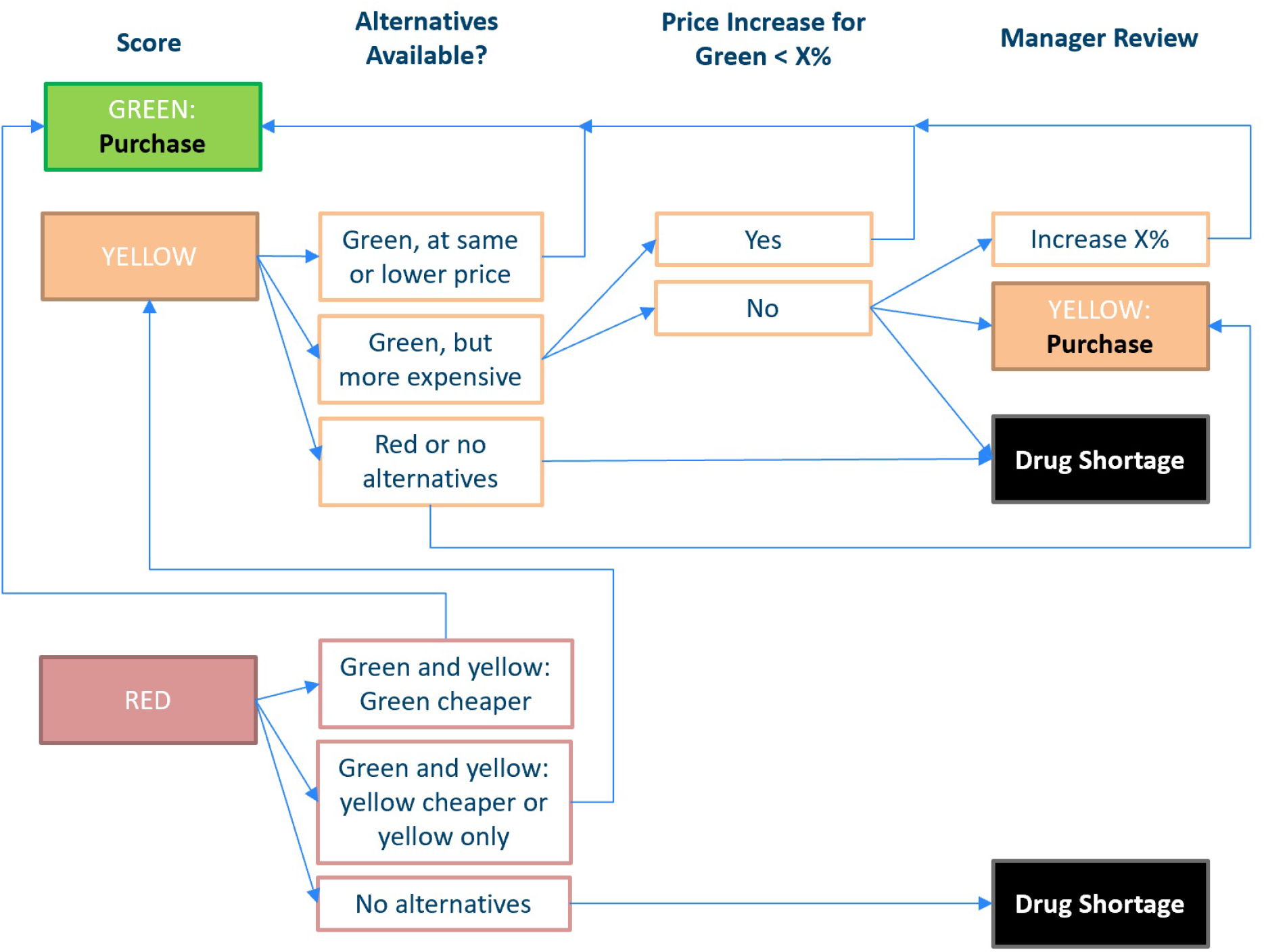
Proposed decision tree implementing red/yellow/green quality score designations. The first column describes the color designation of a drug product that is the default selection for the health system, which then triggers the decision tree.

## Results

The angiotensin receptor blocker drug, valsartan, has been subject to heavy scrutiny over quality due to a multitude of recalls after carcinogenic impurities were found.^23^ This drug has been selected here for analysis using available data to generate a limited number of quality score criteria which give illustrative examples of how such quality scores can be derived. Regulatory information was gathered by Govzilla and chemical analysis information was acquired from Valisure’s analytical laboratory that is attached to a licensed pharmacy.

**Table 4.** Detailed regulatory information (Table 4A) and chemical analysis information (Table 4B) on available manufacturers of valsartan. Although the names have been deidentified, the data describes real manufacturers of valsartan drug products being currently sold in the United States.

| Table 4A | Regulatory Information (2017 – 2020) |  |  |  |  |  |  |
| --- | --- | --- | --- | --- | --- | --- | --- |
|  | Inspections | Form 483 | Warning Letter | Form 483 Ratio | Warning Letter Ratio | Form 483 % Above Global Ratio | Warning Letter % Above Global Ratio |
| Company A | 38 | 20 | 0 | 0.526 | 0.000 |  |  |
| Company B | 6 | 1 | 0 | 0.167 | 0.000 |  |  |
| Company C | 36 | 24 | 1 | 0.667 | 0.028 | 26% |  |
| Company D | 15 | 9 | 1 | 0.600 | 0.067 | 13% | 181% |
| Company E | 10 | 3 | 0 | 0.300 | 0.000 |  |  |
| Company F | 53 | 27 | 0 | 0.509 | 0.000 |  |  |
| Global 3-year | 6,967 | 3,691 | 257 | 0.530 | 0.037 |  |  |

| Table 4B | Chemical Analysis |  |  |  |  |  |  |
| --- | --- | --- | --- | --- | --- | --- | --- |
|  | Batches Analyzed | DMF >96ng, <1000ng | DMF >1000ng | NDMA >96ng | Dosage | Dissolution | Ingredients |
| Company A | 6 | 1 | 0 | 1 | 0 | 0 | 0 |
| Company B | 2 | 2 | 0 | 0 | 0 | 0 | 0 |
| Company C | 9 | 9 | 0 | 0 | 0 | 0 | 0 |
| Company D | 7 | 0 | 0 | 0 | 0 | 0 | 0 |
| Company E | 7 | 0 | 7 | 0 | 0 | 0 | 0 |
| Company F | 2 | 2 | 0 | 0 | 0 | 0 | 0 |

**Table 5.**
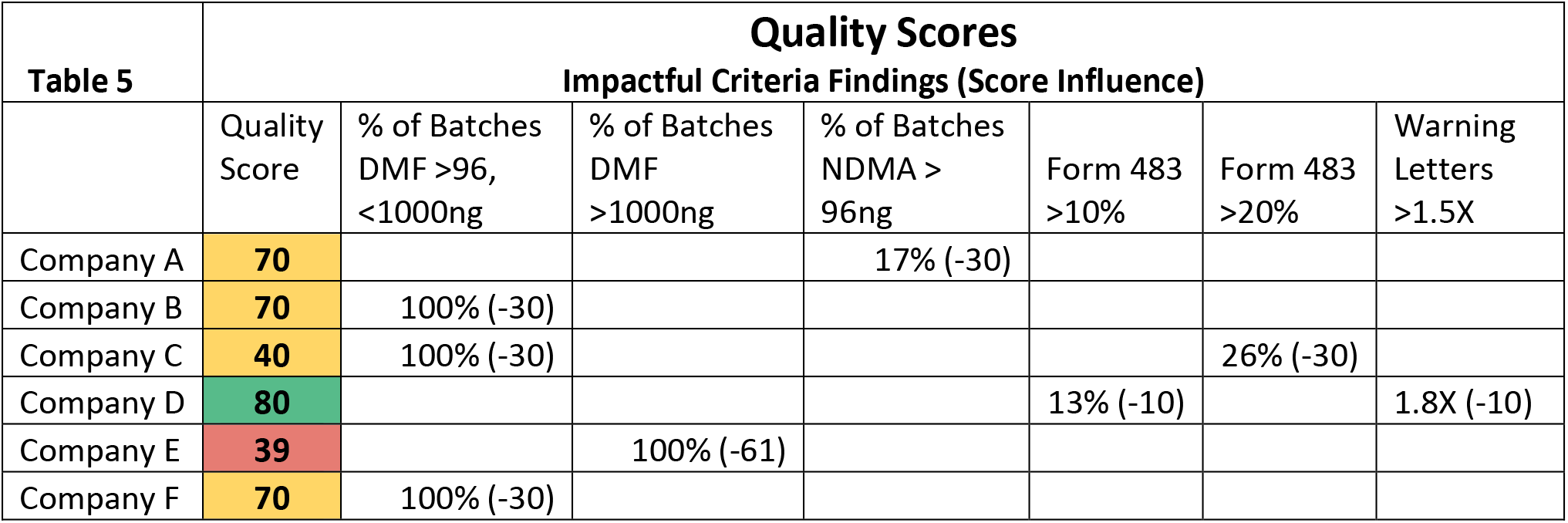
Data output for criteria triggering an influence on quality scores and the corresponding numerical influence on the scores denoted in parentheses, regarding current, on-market valsartan drug products from specific manufacturers. The final calculated quality scores are displayed and given their corresponding color designation.

Even with a drug such as valsartan that has had many quality issues, some of which appear to persist, the use of the proposed quality score system is able to identify a supplier that scores a green. Even among potentially mediocre product quality choices, those that appear to perform particularly poorly are identified by a red and can be reasonably avoided.

To further evaluate the impact on pricing by using the proposed quality score system, the relative costs of the valsartan drug products were analyzed across the six companies. Four dosage forms (40mg, 80mg, 160mg and 320mg) were evaluated using pricing from three different distributors and ensuring packaging size was consistent among all companies.

**Figure 2.**
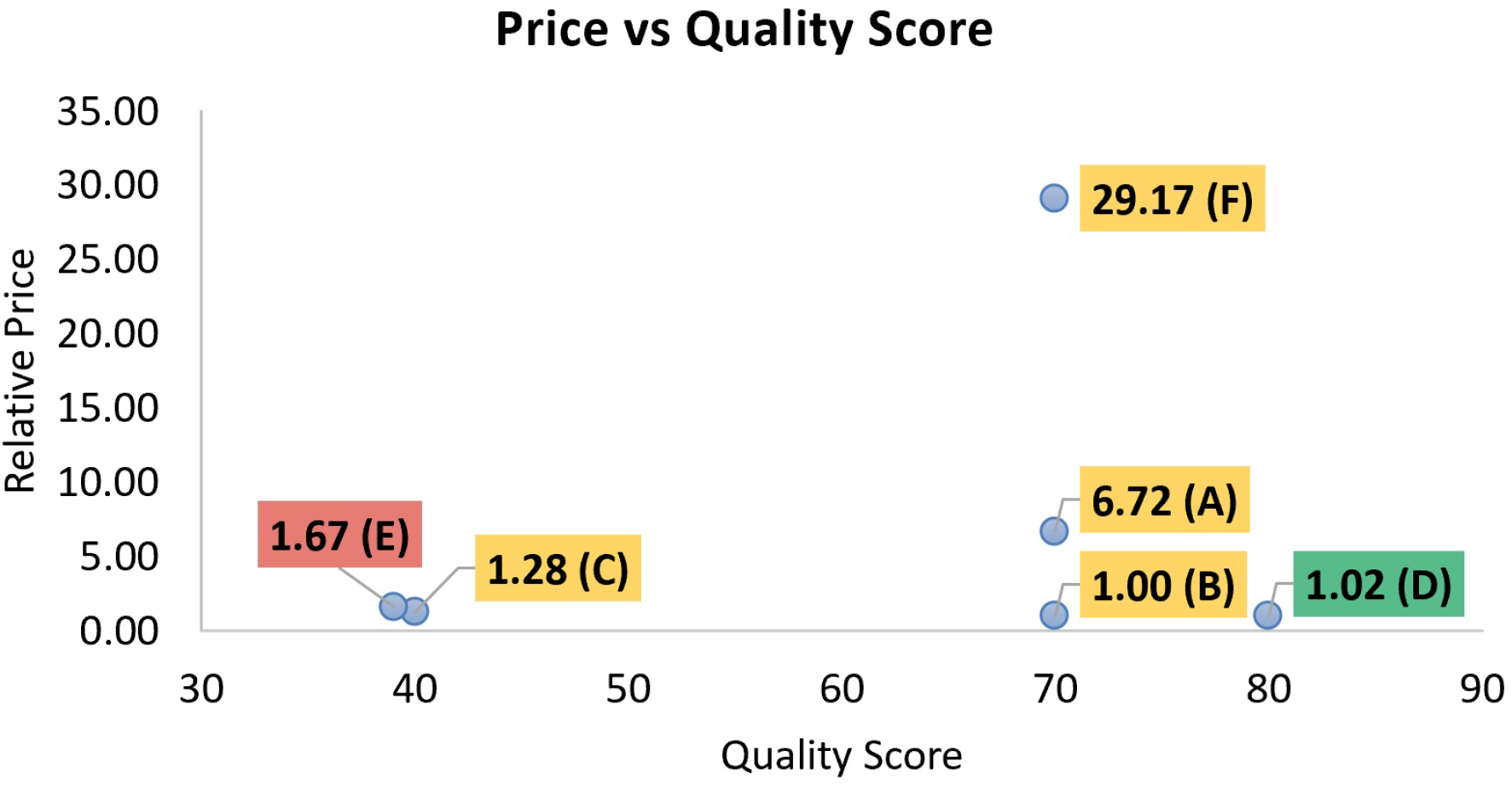
Relative pricing of drug products from companies A – F (denoted in parenthesis) plotted against their quality scores and given their respective red/yellow/green designation.

Although the decision tree in Figure 1 proposes the option of paying more for a higher scoring drug product, the pricing comparison illustrated above suggests that higher quality drug products do not necessarily cost more. Despite continued quality issues with valsartan, the least expensive option had the second-highest quality score, the highest quality score option was only 2% more expensive and the lowest scoring option was 67% *more* expensive.

## Discussion and Conclusion

When originally conceived, generic drug products were assumed to be equal in quality to each other and to the innovator product so the only differentiating feature would be the price paid. This has led to automatic generic substitution laws across the country where patients receive the generic selected by the pharmacy and this could change several times over the patient’s course of therapy. The premise that every innovator and generic product is of equal quality is demonstrably false.^8^

With the changing market dynamics that drove pharmaceutical manufacturing offshore and made it very difficult to warrant acceptable quality, a new strategy is needed to ensure patient safety. The use of drugs that are improperly dosed as well as products that don’t dissolve properly can put patients at risk of clinical failure or adverse events. The use of products with bacterial contamination, unacceptably high amounts of carcinogens or heavy metals may lead to unintended health problems as a result.^24^

We hope this will be a useful overview and baseline proposal for the use of quality scores for drug products. This is critical for adding much-needed transparency into the American drug supply chain and enabling health system purchasers and payers of medications to avoid low-quality drug products. As the data demonstrates with valsartan, high quality drug products do not necessarily cost more. Thus, even if a health system is unable or uninterested to add any additional purchasing cost or add any potential drug shortage burden, it is highly likely that the use of the proposed quality score system will provide a significant benefit in avoiding low-quality drug products.

Such action taken by established healthcare systems could help protect them from recalls and drug shortages while serving as a significant market driver to incentivize the manufacturing industry to produce quality products. Furthermore, the proposed quality score system could provide regulatory agencies with transparent and rational metrics with which to reward high-scoring manufacturers (e.g. faster ANDA approvals) and/or penalize low-scoring manufacturers (e.g. slower and more scrutinized drug approvals).

Overall, drug quality scores have the potential to improve public health; therefore, their continued development and implementation is highly encouraged.

## Data Availability

THere are no patient/human subject data included in this manuscript

## Acknowledgement

The authors would like to thank Michelle Call and Michael de la Torre from Govzilla, Martin Van Trieste from Civica Rx and Amber Jessop, Kaury Kucera and David Light from Valisure, for their assistance in providing data and expertise for this paper.

